# Work-related stress and consumption of psychoactive substances and medications among early childhood professionals in Orléans Métropole, CCTVL, and Fleury-les-Aubrais (TraPsyCOL): Study protocol for a cross-sectional study

**DOI:** 10.64898/2026.02.25.26347115

**Authors:** Walaa Khazaal, Stéphane Onnée, Roomila Naeck, Séverine Morisset-Lopez, Patrick Baril, Olivier Vernay, Raphaël Serreau

## Abstract

Work-related stress is a major public health issue affecting workers across various sectors. Individuals experiencing work-related stress are more likely to consume psychoactive substances, primarily alcohol, tobacco, and cannabis, as well as psychoactive medications, which may be used as coping mechanisms. Work-related stress is also associated with adverse outcomes such as burnout, depression, anxiety, and sleep disorders.

In France, early childhood professionals, including “ATSEMs”, “animateurs”, and “agents d’entretien”, play a crucial role in the education, care, and well-being of children but are exposed to high levels of occupational stress due to the emotionally demanding nature of their work and the associated physical strain, making them vulnerable to substance use, burnout, depression, anxiety, and sleep disorders.

This cross-sectional epidemiological study, conducted at a single time point, will be carried out among early childhood professionals working in schools for children in Orléans Métropole, Communauté de Communes des Terres du Val de Loire (CCTVL), and Fleury-les-Aubrais. Ethical approval for this study was obtained from the Ethics Committee of the Centre Hospitalier Universitaire d’Orléans (assigned reference number is CERO 2511-02).

The study aims to provide a better understanding of the relationship between work-related stress and the use of psychoactive substances and medications among early childhood professionals, as well as the association between work-related stress and burnout, depression, anxiety, and sleep disorders. Data will be collected anonymously using self-administered online questionnaires, accessed via a QR code printed on flyers distributed to participants.

The same QR code will also provide access to an information sheet explaining that the study complies with ethical guidelines and that proceeding implies non-objection to participation. Based on calculations performed using BiostaTGV, a sample size of 265 participants is required. Statistical analysis will be conducted using SPSS software. Studying these associations is essential for informing the development of targeted interventions and prevention.

## I. Introduction

Work-related stress, also known as occupational stress or job stress, is a major alarming public health concern that has been increasing steadily over the past few decades among workers in all sectors. It is defined as “a pattern of reactions that occurs when workers are presented with demands or pressures (stressors) that are not matched to their knowledge, abilities and skills and which challenge their ability to cope” [1]. An estimated 19 to 30% of employees among the general working population are affected by workplace stress and burnout [2, 3]. Work-related stress contributes to 60% of workplace accidents, 40% of turnover, and 19% of absenteeism. Moreover, 10-13% of suicide deaths are due to working conditions. When job demands exceed a person’s resources, needs, or abilities, work stress occurs, resulting in negative psychological and cognitive effects. The most important components of these demands are the workload, physical work environment, difficulties balancing work and life, and emotional strain. The scientific community is giving more attention to job stress because it has an impact on job performance, physical and mental health [4–7].

In our era, a common problem that is experienced by workers in all professions is psychological distress at work, which results from several adverse health conditions related to workers’ health, safety, and wellbeing. Higher levels of psychological distress have been observed among employees in human service occupations like education, social work, and healthcare [2]. According to 2012 research, psychological distress was experienced by 1.4% of men and 3.1% of women in France [8]. A study showed that among a large French company, the prevalence of psychological distress among employees was 32.8% (95 % CI 31.8–33.9 %), higher among women (48.9 %, 95 % CI 46.5–51.7 %) than men (30.1 %, 95 % CI 29.0–31.2 %) [9]. Job strain, as defined by Karasek [10], is the combination of high psychological demands, including task quantity, complexity, and time pressure, and low decision latitude, including both decision authority and skill discretion. It also includes a dimension of social support at work [11]. This encompasses both instrumental and socio-economic support from interactions with managers and colleagues. Insufficient social support in the workplace is considered a potential risk factor for health. In France, a recent study conducted between November 2023 to January 2024 across 104 assisted reproductive technology (ART) centers, reported that among 464 participants including biologists (n = 124), gynecologists (n = 129), technicians (n = 107) secretaries (n = 51), midwives (n = 31), and other professions (n=22), 35.7% of professionals in French ART centers experienced job strain [12]. Another French study conducted in 2016 found that 23% of employees were exposed to job strain [13]. Moreover, data from the SUMER study that was conducted in France, revealed that women (28.2%) were more frequently exposed to job strain, based on the Karasek model, than men (19.6%) [11, 14].

Work-related stress is associated with multiple adverse health consequences and comorbidities, including mental disorders, depression, anxiety [15], sleep disorders [6, 16], and burnout [2]. Burnout is “a syndrome conceptualized as resulting from chronic workplace stress that has not been successfully managed”, and characterized by emotional exhaustion, depersonalization, and reduced personal accomplishment in work. The literature has widely documented the association between job stress and negative mental health outcomes. Workers are more subjected to mental health problems such as depression and anxiety due to stressful work environments. Based on a study done in 2007, people with high psychological demands at work, including both heavy workloads and intense time pressure, were twice as likely to suffer from depression or generalized anxiety disorder than people who had lower job demands. Both women (1.90 [95% CI: 1.22–2.98]) and men (2.00 [95% CI: 1.13–3.56]) showed this association [17]. Another study showed that high job strain was associated with 3.8 (95% CI = 2.8-5.1) times higher odds of depressive symptoms, 1.7 (95% CI = 1.1-2.6) times higher odds of depressive disorders, and 7.4 (95% confidence interval [CI] = 5.6-9.7) times higher odds of burnout than low job strain among employees [18].

A study conducted among 1265 French pharmacists revealed that work-related stress was significantly associated with anxiety (29.16 ± 22.93 vs 52.56 ± 21.55 vs 71.39± 16.0, normal vs suggestive vs indicative scores of anxiety, respectively; p<0.001) and depression (42.77 ± 26.01 vs 68.56 ± 17.82 vs 76.44 ± 14.68, normal vs suggestive vs indicative scores of depression, respectively; p<0.001). The percentage of anxiety or depression indicative scores was higher for participants with a high level of work-related stress. Moreover, this study also revealed that the proportion of sleep disorders was significantly higher among participants suffering from high levels of work-related stress compared to those with low levels of work-related stress (37.6% vs 12.3%, p<0.001) [19]. A study done in America showed that the nature, quantity, mental demand, working hours, and associated stress of work played a fundamental role in insomnia, where approximately 60% of participants identified that either the development or prolongation of their insomnia was primarily caused by work [20]. Furthermore, a Chinese study done among 2116 oilfield workers showed that their sleep quality was poor, and as occupational stress increased, the prevalence of sleep disturbances increased as well [21].

Similarly, the issue of stress at work can be associated with the consumption of psychoactive substances and medications. Psychosocial job strain is associated with a higher risk of substance use disorders, mainly alcohol, tobacco, and cannabis [22]. Several studies have shown that job stress may be linked to increased psychoactive substance use as a coping mechanism to deal with work pressure and difficulties and to minimize the adverse effects of different sources of suffering, and this helps individuals to relax from job stress [7, 23]. For example, a study conducted using data from the French CONSTANCES cohort shows that men experiencing work-related stress were more likely to consume tobacco (OR=1.17, 95% CI=[1.06;1.29], p=0.002 for former smokers), use cannabis (OR=1.17, 95% CI=[1.06;1.29], p=0.001 for use more than a year ago), and have episodes of excessive alcohol consumption (OR=1.29, 95% CI=[1.10;1.51], p=0.002) compared to those who were not stressed. Exposure to work-related stress was associated with a higher level of tobacco use (OR=1.75, 95% CI=[1.34;2.30], p<0.001), alcohol consumption (chronic use: OR=1.59, 95% CI=[1.12;2.25], p=0.009; dependence: OR=2.30, 95% CI=[1.54;3.44], p<0.001), and cannabis use (OR=1.38, 95% CI=[1.10;1.75], p=0.006 for use more than once a month) compared to those not exposed among women [22].

A recent Brazilian study conducted among university professors revealed a significant association between occupational stress and consumption of psychoactive substances, where among professors experiencing stress at work, 85.7%, 28.6%, and 20% consume alcohol, tobacco, and cannabis, respectively [24]. According to a meta-analysis of European studies, smokers are more likely than non-smokers to report work-related stress (OR=1.11, 95% CI= [1.03;1.18]), and among smokers, those who reported work-related stress smoked on average more cigarettes per week than non-stressed smokers (an average of 3 extra cigarettes, i.e., 103.3 for stressed smokers versus 99.9 for non-stressed smokers) [25]. Moreover, a Canadian study showed that alcohol consumption tends to be higher when the perception of mental health and work-related or life stress is low. This study highlights the role that the work environment could play in preventing stressful factors and reducing alcohol consumption [26]. In Spain, researchers have reported a doubling of the risk of increased alcohol consumption based on exposure to a harmful work environment (OR=2.15, 95% CI=[1.51;3.06] for moderate exposure level, OR=2.23, 95% CI=[1.49;3.36] for high exposure level) [27]. Additionally, the study of Balayssac et al. (2017) demonstrated that among 421 French male pharmacists, 16.7% of those with a high level of stress at work had alcohol consumption above the WHO limit, compared to 4% of men whose consumption was below the World Health Organization (WHO) limit (p < 0.001) [19].

Furthermore, the study of CONSTANCES data highlights the effect of public contact on substance consumption: the consumption of tobacco and cannabis among professionals in contact with the public was higher than among professionals without public contact, for both men (OR=1.47, 95% CI=[1.22; 1.78], p<0.001 for high cigarette consumption; OR=1.38, 95% CI=[1.17; 1.63], p<0.001 for cannabis consumption, less than once a month) and women (OR=1.63, 95% CI=[1.18; 2.25], p=0.003 for high cigarette consumption; OR=1.31, 95% CI=[1.05; 1.63], p=0.017 for cannabis consumption, less than once a month). Alcohol consumption (chronic, at risk, or significant occasional) was higher among men in contact with the public compared with those who were not (OR=1.71, 95% CI=[1.43; 2.05], p<0.001 for moderate chronic consumption; OR=1.22, 95% CI=[1.12; 1.33], p<0.001 for at-risk consumption; OR=1.33, 95% CI=[1.18; 1.51], p<0.001 for significant occasional alcohol use more than once a month) [22].

Regarding the association between work-related stress and the consumption of psychoactive medications, a meta-analysis of 18 unique studies showed that high job demands were associated with a statistically significant increased risk of psychotropic medication use (RR 1.16, 95% CI 1.02 to 1.31). In studies reporting OR, high job demands were associated with an OR of 1.39 (95% CI 1.06 to 1.71) [28]. Another study conducted among French community pharmacies showed that anxiolytic and hypnotic drug use was significantly associated with work-related stress scores, where anxiolytic and hypnotic drug users were more stressed (score ≥70) than non-users (anxiolytics: 65.1% *vs* 34.9%, *p* < 0.001; hypnotics: 66.7% *vs* 33.3%, *p* < 0.001) and had higher stress scores (anxiolytics: 71.97 ± 17.34 *vs* 48.75 ± 27.19, *p* < 0.001; hypnotics: 73.35 ± 19.74 *vs* 51.15 ± 27.12, *p* < 0.001) [19].

In France, early childhood professionals, including “Agents Territoriaux Spécialisés des Écoles Maternelles (ATSEMs)”, “animateurs”, and “agents d’entretien”, play an important role in the well-being of children. “ATSEMs” are workers who help teachers in preschool classes; they are responsible for welcoming children, ensuring their hygiene, providing educational and technical support in the classroom, helping teachers with teaching staff during school hours, preparing materials, maintaining the class environment, as well as managing and assisting children throughout extracurricular time, such as breaks for lunch and care after school [29]. Early childhood “animateurs” are workers that play an important role in supporting the education, well-being, and development of children during extracurricular periods only outside regular school hours, such as the time before classes starts, during children’s meal and nap periods, and after school hours. They are implicated in organizing the children’s daily routine within the school, in the leisure centers, and during vacation. They also play with the children, supervise them in the playground, and organize and manage safe educational, artistic, and recreational (such as games and sports) workshops based on their needs, which aids in promoting the development, learning, and socialization of young children [30]. “Agents d’entretien” in charge of early childhood professionals are workers that maintain the safety and hygiene of children; they are responsible for cleaning and sanitizing areas such as floors, bathrooms, and play areas efficiently and effectively to ensure the health and safety of the children [31].

The working conditions of early childhood professionals expose them to high levels of psychosocial stress and burnout. Indeed, this profession face challenging working conditions, including material and human constraints, lack of recognition, paradoxical demands, exhaustion caused by children’s crying and screaming, and more. They often work long hours, manage large groups of young children, deal with the emotional needs of children, and interact with parents. Early childhood educators (ECEs) face many stressors through their work, such as high workload, stressful environment, limited chances of progress, and low pay, which cause low levels of job satisfaction and higher rates of turnover, as well as negative impacts on themselves, the field, and the children they work with [32]. A recent study conducted among 137 ECEs showed that 62% suffer from workload and 44% from job strain, compared to 35% and 36%, respectively, within the national sample [33].

In France, a recent study conducted in 340 nursery schools by the CNRACL National Prevention Fund among ATSEMs, which had a 67% response rate from 1,347 questionnaires distributed, showed that 62% think that their work negatively impacts their physical or psychological health, 82% stated that their daily activities induce more anxiety, and half of the respondents feel a deterioration in their sleep quality due to their work. Moreover, this study showed that this population suffers from a lack of support from the community and feelings of isolation [34]. Similarly, according to an in-depth study on childcare assistants, the average rate of long-term sick leave (90 days in the last 12 months) due to psychosocial risks in this population exceeds that of other territorial professions (30% compared to 25% for other professions) [35]. This could reflect dissatisfaction within this profession, despite its key role in preventing potential risky behaviors (addictions, dangerous behaviors, etc.) that children may develop later on. In fact, according to a study by the Rhône-Alpes Regional Health Observatory, early childhood professionals are “hidden actors” in this prevention. Therefore, their health status, especially mental health, at work is a major public health issue [36].

Early childhood professionals may also be affected by depressive disorders, with possible consequences on the behavior of children who are sensitive to the emotions of those around them [37]. The study of Farewell et al. (2022) demonstrated that among 136 Early Childhood Educators, the rates of depression were 2.3 times higher than nationally available normative data [33], which were consistent with the study of Linnan et al. (2017) showing that the rates of depression among this population are three times the national average (36% vs. 12% clinically depressed) [38].

Investigating the relationship between work-related stress and addictions among professionals outside the healthcare sector would be enriching, especially regarding early childhood professionals, whose role is crucial, and whose working conditions are challenging. This unique combination of stressors makes them an important population for studying the association between workplace stress and substance use. Despite their essential role, no studies have yet examined this association among early childhood professionals in France, particularly in Orléans Métropole, CCTVL, and Fleury-Les-Aubrais, making this the first investigation of its kind in these territories. We hypothesize that work-related stress is associated with the consumption of psychoactive substances and medications, as well as addiction, among early childhood professionals (“ATSEM”, “animateurs”, and “agents d’entretien”), and such consumption serves as a coping mechanism for occupational stress.

By focusing on early childhood professionals, this study has the potential to inform the development of targeted interventions and workplace policies aimed at reducing stress and preventing substance use in this vulnerable group, and to better target professionals at risk. This could have significant implications for their well-being and overall job satisfaction, as well as for the well-being of the children under their care.

## II. Materials and methods

### II.1 Aims

#### II.1.1 Primary aim

To assess the association between work-related stress and the consumption of psychoactive substances (alcohol, tobacco, and cannabis) and medications among early childhood professionals working in schools for children in Orléans Métropole, CCTVL, and Fleury-les-Aubrais, and to evaluate the level of dependence on these substances.

#### II.1.2 Secondary aims

- To evaluate the association between work-related stress and burnout.
- To assess the relationship between work-related stress and anxiety and depression symptoms.
- To study the association between work-related stress and sleep disorders.

### II.2 Study design

This study is a multicenter study, observational, cross-sectional epidemiological study conducted in Orléans Métropole, CCTVL, and Fleury-les-Aubrais. It is a non-interventional study with minimal risk and constraints, involving human participants.

The study population consists of early childhood professionals, mainly “ATSEMs”, “animateurs”, and “agents d’entretien”, working in schools for children within the aforementioned areas.

### II.3 Sample size

The minimum required sample size was calculated using the BiostaTGV, an online biostatistics platform. Based on the practical experience of Pr. Raphael SERREAU, who worked as an addictologist and occupational physician in the regions where the study will be conducted, between 2013 and 2025, it was observed that among the 7,700 agents he followed, approximately 30% of early childhood professionals were addicted to at least one type of psychoactive substances or medication.

Assuming a prevalence of psychoactive substances or medication use of 30%, and assuming an odds ratio (OR) of 2.4, a power of 80% and an alpha of 0.05 (5%), the minimum sample size calculated for our study was estimated at 238 participants. To account for an estimated 10% loss due to incomplete questionnaires or missing data, a total of 265 participants will be included.

This sample size represents a significant proportion of the target population in Orléans Métropole, CCTVL, and Fleury-les-Aubrais municipalities (more than 600 early childhood professionals in Orléans Métropole, more than 200 in Fleury-Les-Aubrais, and more than 70 in CCTVL), and is considered sufficient for identifying statistical associations between work-related stress and the use of psychoactive substances or medications, as well as related mental health outcomes, within a cross-sectional study design.

### II.4 Eligibility criteria

#### II.4.1 Inclusion criteria

Participants will be eligible for inclusion if they meet all of the following criteria:

- Age ≥ 18 years;
- Currently employed as an “ATSEM”, “animateur”, or “agent d’entretien” working in early childhood settings in Orléans Métropole, CCTVL and Fleury-les-Aubrais;
- Employees who have been working in their current position for at least 12 months at the time of study inclusion;
- Employees who voluntarily agree to participate in the study after reading the information sheet and express their non-opposition by completing the questionnaire.

#### II.4.2 Non-inclusion criteria

- Employees who decline participation in the study.

#### II.4.3 Exclusion criteria

- People who initially agreed to share their data but later changed their minds and no longer wish to do so.

### II.5 Participant characteristics and sampling procedure

The study population consists of early childhood professionals, specifically “Agent Territoriaux Spécialisé des Écoles Maternelles” (ATSEM), “animateurs”, and “agents d’entretien” employed in schools for children in Orléans Métropole, CCTVL, and Fleury-les-Aubrais.

Information about the study will be communicated to early childhood professionals (“ATSEMs”, “animateurs”, and “agents d’entretien”) through the directors and the research team (Ms. KHAZAAL, Pr. SERREAU, and Pr. ONNEE). The research team will be available on-site to answer questions and provide further clarifications as needed. In addition, informational flyers presenting the study objectives and significance will be distributed to all staff.

Inclusion will be based on voluntary participation following the presentation of the study in detail and the distribution of informational flyers. All eligible professionals present in the selected structures at the time of data collection will be invited to participate in the study. To ensure representativeness and protect anonymity, all staff members, regardless of their intention to participate, will be approached equally, and flyers will be distributed to the entire staff. The flyers will include a QR code that, once scanned, links to an information sheet explaining that the study complies with ethical guidelines, informs participants of their rights, and states that by continuing, they do not object to participating. The same QR code will also link directly to the online self-administered questionnaire following the information sheet. The study is completely anonymous, and no personal identifying information (e.g., names, phone numbers, or emails) will be collected from participants.

### II.6 Study procedure

This study is a multicenter observational cross-sectional epidemiological study that will be conducted in Orléans Métropole, CCTVL, and Fleury-les-Aubrais regions.

The objective of the study is to assess the relationship between work-related stress and the use of psychoactive substances (alcohol, tobacco, and cannabis) and medications, as well as the relationship of work-related stress with burnout, depression, anxiety, and sleep disorders. Official approval was obtained from the heads of the three regions, following meetings held with the Directors General of Services from those regions in 2025. Moreover, ethical approval was obtained from the Ethics Committee of the Centre Hospitalier Universitaire (CHU) d’Orléans on 5 December 2025 (assigned reference number: CERO 2511-02).

Self-administered online questionnaires accessed via QR codes will be used for data collection, and will be completed by participants. The online questionnaire will take approximately 30–40 minutes to complete. Creative flyers including the study title, objectives, and significance will be created to motivate the targeted population to participate. Each flyer will include a QR code linking directly to the online questionnaire accessible through participants’ phones. In addition to the questionnaire, the same QR code will link to an information sheet presented to participants immediately before the questionnaire. This sheet will explain that our study complies with ethical guidelines, inform participants of their rights, and state that by continuing, they do not object to participating. It will also provide the investigator’s (Ms. Walaa KHAZAAL) email address for any questions.

Prior to the start of data collection, meetings will be held separately with stakeholders and educational directors of schools in each study region (Orléans Métropole, CCTVL, and Fleury-les-Aubrais). The purpose of these meetings is to present the study in detail, including its objectives, methodology, procedures, and overall relevance to the targeted professions.

As educational directors are familiar with the working environment and are in direct contact with early childhood professionals, discussions during these meetings will aim to identify any practical constraints related to the implementation of the study and to ensure that the questionnaires and study procedures are understandable and feasible in the field. This step is intended to allow form some adjustments to the questionnaire or study procedures, if necessary, based on their experience before the launch of the quantitative data collection.

These discussions will also aim to obtain the educational directors’ agreement to facilitate the implementation of the study and to jointly define together a clear and feasible plan for how the data collection will be carried out. As previously discussed and agreed with representatives in Orléans Métropole, CCTVL, and Fleury-les-Aubrais, once the directors have a clear understanding of the study, they will help the research team by informing staff members (“ATSEMs”, “animateurs”, “agents d’entretien”) who will be gathered on a dedicated day about the study and the importance of their participation. They will also assist, together with the research team, in distributing flyers containing the QR codes, which will include the investigator’s email address, allowing participants to contact her privately and confidentially if they have questions or require further clarification before they complete the questionnaires or decide whether to participate.

### II.7 Data collection process

After the meetings with the directors, and on a specific day coordinated with each director, all staff (“ATSEMs”, “animateurs”, “agents d’entretien”) will be gathered and informed about the study and its details by their directors. The research team will also be present to answer any questions and provide further explanations and clarifications as needed. Flyers containing the QR codes will be distributed to all professionals by the directors and the research team. Participants will be able to scan the QR code, using their phones, which is linked directly to the online questionnaire at a time that suits them, either during working hours or outside of work, within a frame time of one month, with the expectation that many participants will respond on the same day it is presented while the information is still fresh and clear. During the one-month period, educational directors will send general reminders to all staff members, encouraging those who have not yet completed the questionnaire to do so within the designated timeframe.

No identifiable personal information such as names, emails, or phone numbers will be collected to preserve full anonymity of participants, and to allow them to feel more comfortable upon answering sensitive questions regarding work-related stress, the use of psychoactive substances and medications, and their mental health without fear of identification. Participants will also be assured that their data will remain confidential and that participation is entirely voluntary. Participants will complete the online questionnaires either during or outside of working hours, within a timeframe of one month, taking approximately 30–40 minutes.

The questionnaire used for data collection was composed of several parts:

1. Sociodemographic and socio-economic characteristics: age, gender, marital status, having children, educational level, and household composition.
2. Work-related variables: professional role, working hours, job income, duration of employment, job satisfaction, perceived value of their work, relationship with children’s parents, and relationship with teachers.
3. Assessment of work-related stress: The Karasek’s Job Content Questionnaire (JCQ) will be used to assess work-related stress. Participants will be classified into an exposed group (experiencing work-related stress), defined by high psychological demands and low decision latitude, and a non-exposed group (not experiencing work-related stress), defined by other combinations of psychological demands and decision latitude.
4. Assessment of outcomes associated with work-related stress: A battery of validated French-language instruments will be used, including Fagerström Test for Nicotine Dependence (FTND), Alcohol Use Disorder Identification Test (AUDIT), Cannabis Abuse Screening Test (CAST), and the Alcohol, Smoking and Substance Involvement Screening Test (ASSIST) for psychoactive medication use. Burnout will be assessed using the Maslach Burnout Inventory (MBI), symptoms of depression and anxiety using the Hospital Anxiety and Depression Scale (HADS), and sleep disorders using the Epworth Sleepiness Scale.

### II.8 Data Measurements

#### II.8.1 Karasek’s Job Content Questionnaire

The Job Content Questionnaire (JCQ), developed by Karasek, is an international self-assessment questionnaire used to assess the level of psychosocial stress at work. It is the main instrument used to evaluate three dimensions of the work environment: the psychosocial demands (9 items), the decision latitude, (9 items) and the social support (8 items). The questionnaire includes 26 items with a 4-point Likert scale ranging from strongly disagree (1) to strongly agree (4).

The three dimensions are calculated through the three following formulas [10, 39]:

1. psychological demands (Q10 + Q11 + Q12 + (5 − Q13) + Q14 + Q15 + Q16 + Q17 + Q18), where a score below 20 refers to low psychological demands,
2. decision latitude (4* Q4 + 4* (5 − Q6) + 4* Q8 + 2* (5 − Q2) + 2* Q5 + 2* Q7 + 2* Q1 + 2* Q3 + 2* Q9), where a score below 71 refers to low decision latitude,
3. social support (Q19 + Q20 + Q21 + Q22 + Q23 + Q24 + Q25 + Q26), where a score below 24 reflects low social support.

The combination of psychological demand and decision latitude can give four situations at work:

- Relaxed work (low demand and high decision latitude),
- Passive work (low demand and low decision latitude),
- Active work (high demand and high decision latitude),
- Stressed, tense work (high demand and low decision latitude).

Job strain is defined as a psychological demand score higher than 21 and a decision latitude score lower than 70. A social support level < 24 indicates isostrains [40]. In this study, we will use the validated French version of the JCQ [41].

#### II.8.2 Fagerström Test for Nicotine Dependence (FTND)

This test is used to assess the level of nicotine dependence among current smokers [42]. It consists of 6 items, which are summed to give a total score ranging from 0 to 10. Nicotine dependence is categorized into five levels: no dependence (0 to 2), low dependence (3 to 4), medium dependence (5 to 6), high dependence (7 to 8) and very high dependence (9 to 10). The validated French version of the Fagerström Test for Nicotine Dependence will be used in this study [43].

#### II.8.3 Alcohol Use Disorder Identification Test (AUDIT)

This test is a 10-item screening tool developed by the WHO (WHO) to assess alcohol consumption, alcohol-related behaviors, and alcohol-related problems [44]. When the items are summed, they give a score ranging from 0 to 40, which provides an indication of alcohol use disorder severity. Scores between 8 (7 for women) and 15 suggest hazardous alcohol consumption, 16 to 19 harmful alcohol consumption, and > 20 alcohol dependence [45]. We will use the French validated version of AUDIT in this study [46].

#### II.8.4 Cannabis Abuse Screening Test (CAST)

This test is a 6-item screening tool used to assess cannabis-related problems [47]. The response options are rated on 5-point Likert scale ranging from 0 (never) to 4 (very often). The total score obtained after summing the items can range from 0 to 24 and indicates whether or not the respondents are at risk. A score less than 3 indicates no addiction risk, a score between 3 and 6 indicates low addiction risk, and a score of 7 or above indicates high addiction risk [48]. The French version of CAST will be used in this study [49].

#### II.8.5 The Alcohol, Smoking and Substance Involvement Screening Test (ASSIST)

This reliable instrument, developed by the World Health Organization, is designed to identify problematic or risky substance use, including substances used without medical supervision as well as the non-medical use or misuse of prescribed medications.

It includes eight core questions applied across multiple substance categories (tobacco, alcohol, cannabis, cocaine, amphetamine-type stimulants, inhalants, sedatives, hallucinogens, opiates, and other drugs). In the present study, the ASSIST will be administered only for substance categories other than tobacco, alcohol, and cannabis, as validated instruments (AUDIT, FTND, and CAST) are already used to assess these substances. The instrument will focus on substances used for reasons other than medical prescription, or more frequently or at higher doses than those recommended.

Question 1 assesses lifetime use of substances, while Question 2 addresses the frequency of substance use during the past three months. Responses are rated on a 5-point frequency scale ranging from “never (in the past 3 months)” to “daily or almost daily.” If none of the substances have been used in the past 3 months, the interviewer skips directly to the last three questions concerning lifetime problems and patterns of former use. If any substance has been used during the past 3 months, Questions 3–5 are asked before concluding with Questions 6–8.

Question 3 explores the compulsion to use substances in the previous 3 months (a measure of psychological dependence). Question 4 screens for health, social, financial, or legal problems related to substance use within the past 3 months. Question 5 investigates whether participants have failed to meet role obligations. Questions 6–8 screen for lifetime and recent problems, including concerns from relatives, previous attempts to control substance use, and current or lifetime drug injection. For each substance, a score between 0 and 3 indicates non-problematic use (low risk), a score between 4 and 26 indicates abuse (moderate risk), and a score of 27 or higher indicates dependence (high risk). We will use the validated French version of the ASSIST questionnaire [50, 51].

#### II.8.6 Maslach Burnout Inventory (MBI)

The prevalence of burnout is the main outcome of this scale. We will use the validated French version of the Maslach Burnout Inventory – Human Services Survey (MBI - HSS) in our study [52, 53]. It consists of 22 items, each of which is scored from 0 (never) to 7 (several times a day). 9 of the items evaluate emotional exhaustion, 5 evaluate depersonalization, and 8 evaluate reduced personal accomplishment. The cut-off score for emotional exhaustion is ≥ 30 and that of depersonalization domain ≥ 12. The cut-off for reduced personal accomplishment is ≤ 33. Burnout is experienced by participants when the level of emotional exhaustion or depersonalization exceeds the cut-off, regardless of the presence or absence of reduced personal accomplishment [54, 55].

#### II.8.7 Hospital Anxiety and Depression Scale (HADS)

This 14-item (7 items for each subscale: depression and anxiety) self-report scale is used to assess anxiety and depression and their severity [56]. We will use the French version of the HADS questionnaire, which has demonstrated a reliable and valid clinical assessment of depression and anxiety [57]. Each item is scored on a 4-point Likert scale. The total score of each subscale is obtained by summing the respective 7 items, ranging from 0 to 21. Three severity ranges based on cutoff scores are used: 0–7 (non-cases), 8–10 (mild severity), and 11–21 (moderate or severe severity) [58].

#### II.8.8 Epworth Sleepiness Scale (ESS)

It is an 8-item questionnaire used to assess the level of daytime sleepiness, which is associated with various sleep disorders. The validated French ESS scale version will be used in this study. The items of the scale have a four-point scale, where ‘0’ indicates ‘would never nod off’, while ‘3’ indicates a ‘strong chance of nodding off’. The scale questions refer to eight different situations encountered in daily life. The ESS total score is generated by adding all the individual item scores, ranging between 0 and 24. A score of 10 or less indicates no excessive daytime sleepiness, while a score greater than 10 indicates excessive daytime sleepiness [59, 60].

### II.9 Data management

Data management will include secure collection, storage, and analysis of data, with measures to ensure confidentiality and participant anonymity. The data will be stored and will be only accessible to Ms. KHAZAAL. The results will be analyzed and reported in aggregate form. Data cleaning will be included in the data management process to ensure the quality and consistency of the data. Data management will be supervised by the Centre Hospitalier Universitaire (CHU) d’Orléans team dedicated to clinical research and statistical analysis.

### II.10 Safety considerations

This study is a non-interventional observational study involving minimal risk and constraints. Participation is entirely voluntary and anonymous. Participants may stop completing the questionnaire at any time without providing justification and without any consequences. No specific procedures are required in the event of withdrawal.

In accordance with applicable legislative and regulatory provisions, in particular articles L.1121-3 and R.5121-13 of the French Public Health Code (CSP), all necessary precautions will be taken to ensure the confidentiality of information relating to the research, the participants and their identities, as well as the results obtained. Only the investigator and authorized members of the research team will have access to the anonymized dataset for quality control and statistical analysis.

To ensure anonymity, the online data collection platform used will not collect any identifiable personal information such as names or phone numbers. Data will be collected electronically using the LimeSurvey digital platform, a secure system designed for confidential data collection and storage, and will then be exported in a compatible format (e.g., CSV or Excel) and analyzed. Digitally collected data will be securely stored and protected, with access restricted to the research team. In accordance with Article R.1123-61 of the CSP, the investigator will retain the research-related documents and anonymized data for at least 15 years after the end of the study or its early termination, without prejudice to applicable legislative and regulatory provisions.

Information regarding participants’ rights, data confidentiality, and ethical compliance will be clearly explained in the information sheet, which participants will access via QR codes prior to completing the questionnaire.

### II.11 Statistical Analysis

At the end of data collection, adequate data management will be performed to obtain a clean database. At this stage, database lock will be performed before statistical analysis. The data entry and statistical analysis will be performed using IBM SPSS version 27.

A descriptive analysis of the study population will be conducted. Results will be presented as percentages and frequencies for categorical variables. Graphical representations will be used for better visualization of the data. Prevalence rates of substance use will be calculated. An indicator of work-related stress will be constructed using the JCQ questionnaire based on the collected data.The association between work-related stress and the consumption of tobacco, alcohol, cannabis, and psychoactive medications will be studied separately as well as its association with burnout, depression, anxiety, and sleep disorders.

Bivariate statistical analysis will be performed using the Pearson chi-square test (or Fisher’s exact test) for categorical variables to study the association between each of the relevant independent variables and each of the dependent variables.

Then, multivariable analysis using logistic regression will be carried out for categorical dependent variables in order to detect their correlates. Independent variables that show significant association in the bivariate analysis (p-value<0.2), will be included in multivariable analysis models. The first type error α will be fixed at 5%, and a p-value<0.05 will be considered significant. Results will be presented as adjusted odds ratios (ORa) with their 95% confidence intervals (CI).

### II.12 Ethical considerations and declarations

This study fully complies with current ethical standards for non-interventional research conducted in France.

Ethical approval for this study was obtained from the Ethics Committee of the Centre Hospitalier Universitaire (CHU) d’Orléans (the assigned reference number is CERO 2511-02).

To ensure transparency and adherence to ethical guidelines, the QR code will first link to an information sheet presented to participants directly before the questionnaire. This sheet will explain that our study adheres to ethical guidelines, inform participants about their rights, and state that by continuing, they do not object to participating in the study and proceeding to the questionnaire will be considered as implied consent (non-opposition). It will also include the investigator’s email address in case participants have questions. As the questionnaire is anonymous, withdrawal is possible at any time before submission of the responses.

Each participant will receive detailed information about the purpose, methods, benefits, risks, and confidentiality of the study through communication by the directors and the research team.

Participation will be entirely voluntary and based on a free decision. Questionnaires will be completed anonymously, without collection of participants’ names. The anonymity and confidentiality of collected information will also be explained to participants, a step that remains primary and essential for their motivation and comfort to participate. Data will be collected via a secure online platform and stored safely, and only Ms. KHAZAAL will have access to the data. All data will be used exclusively for scientific purposes, and will be destroyed after the legal retention period, in accordance with CNIL and GDPR regulations.

### II.13 Status and Timeline of the Study

At the current time, official approval has been obtained from the heads of Orléans Métropole, CCTVL, and Fleury-les-Aubrais, following meetings held in 2025 with the Directors General of Services of the three regions. Ethical approval for this study has also been granted by the Ethics Committee of the Centre Hospitalier Universitaire (CHU) of Orléans, following a formal meeting during which the study was presented and discussed. The questionnaire and the participant information sheet have been finalized.

In parallel, preliminary meetings are being conducted with stakeholders and educational directors from schools across the three territories. During these meetings, the research team (Ms. KHAZAAL, Pr. SERREAU, and Pr. ONNEE) provides a detailed presentation of the study, including its objectives, methodology, procedures, and scientific relevance to the target professions. All questions raised by stakeholders are addressed, and any necessary clarifications are provided. These discussions aim to identify potential practical constraints related to study implementation, as the directors are familiar with the working environment and are in direct contact with early childhood professionals. They also help ensure that the questionnaires and study procedures are understandable and feasible in the field. As a result, adjustments to the questionnaire and study procedures have been made before the study is launched. In addition, coordination discussions are being held regarding how we will collect the data and the procedure we will use, and as agreed, educational directors will support the conduct of the study by organizing a dedicated session during which all early childhood professionals will be invited to attend an occasion for addiction. During this session, we will explain the study in detail, encourage them to participate, assure them that the study is completely anonymous, and distribute the flyers containing the QR codes with the directors’ assistance. Some educational directors have already begun informing staff about the study, making it easier to start data collection.

Participant recruitment and data collection is planned to begin in 25 February 2026 and will be conducted across three geographical regions. Due to local administrative procedures, initiation dates may vary slightly between regions. In each region, recruitment and data collection will start simultaneously, as participants will have immediate access to the online questionnaire upon study presentation. In each region, data collection will remain open for one month following the start of recruitment.

Recruitment is expected to be completed by end of April 2026, depending on the different regional start dates. Overall data collection is expected to be completed between end of April and end of May 2026, once all regions have finalized their respective one-month data collection periods. Data collected via LimeSurvey will be exported to an adequate platform, which could be Excel or CSV, after which statistical analysis will be conducted using SPSS software upon completion of data collection. Data management and cleaning will begin immediately after the completion of data collection. Data analysis is expected to be completed by December 2026.

### II.14 Expected results

Participants exposed to job strain are expected to show higher levels of substance use such as alcohol, tobacco, cannabis, and psychoactive medications compared to non-exposed participants. In addition, exposure to job strain is expected to be associated with higher levels of burnout, depression, anxiety, and sleep disorders.

## III. Discussion

To our knowledge, this cross-sectional study is the first conducted in France among early childhood professionals. It sheds light on a population that has not been specifically targeted in previous studies, thereby addressing an important gap in the literature. This study aims to elucidate the relationship between work-related stress and the use of psychoactive substances and medications among early childhood professionals, as well as the association of work-related stress and burnout, depression, anxiety, and sleep disorders.

### III.1. Strengths

From a scientific perspective, this study will provide new epidemiological data on the associations between work-related stress, burnout, mental health disorders (anxiety and depression), sleep disorders, and the use of psychoactive substance and medication use among early childhood professionals, a population that remains underrepresented in the French literature. These findings will contribute to a better understanding of psychosocial risks in the early childhood education sector and may serve as a basis for future interventional or preventive studies.

From a societal and occupational health perspective, the results may inform the development of targeted interventions and workplace policies aimed at improving mental health and well-being among early childhood professionals. The study may also contribute to the design of support systems and practices designed to improve quality of life at work.

The study is completely anonymous and confidential; no personal information will be obtained, and data will be collected digitally using QR codes, which may enhance participation.

The study may provide participants with information regarding their levels of work-related stress and its associations with psychoactive substance and medication uses, as well as with burnout, depression, anxiety, and sleep disorders, thereby helping to improve understanding of these relationships. The study may also contribute to increased awareness of mental health and well-being among participants, highlighting the impact of work-related stress and potentially facilitating access to appropriate support resources.

### III.2. Limitations

Despite the strengths of this study, there are a few limitations. First, the moderate sample size may limit the statistical power of the analyses. Second, the use of a self-administered online questionnaire may introduce information bias, including recall bias and social desirability bias, particularly given the sensitive nature of substance use and psychological distress. However, the fully anonymous and confidential design of the study, with no collection of identifying data and digital data collection through QR codes on the flyers, is expected to encourage honest responses and help reduce these biases; this represents a key strength of this study. Third, the study is conducted in a specific geographical area and targets a specific occupational population, early childhood professionals, which may limits the generalizability of the results to other regions or populations.

### III.2. Dissemination of results

The global findings of the study will be communicated after the completion of data collection, analysis, and reporting. The results will be published in open-access scientific journals, ensuring that all interested parties, including participating centers and individuals, can access them freely. In addition, the study and its results will be presented at national and international scientific conferences in the fields of addiction and mental health, building on the dissemination already initiated through presentations at three conferences to date: BIOTECHNOSCIENCES 2024 Congress (poster presentation) in Nouan-le-Fuzelier, France; the ALBATROS 2025 Congress (oral presentation) in Paris, France; and the C³PO – Clinical Research, Projects, and Precision in Psychiatry 2025 conference (oral presentation) in Orléans, France.

### III.3 Amendments

In the case of any amendments to the study protocol, the revised version will be submitted to the ethics committee of the Centre Hospitalier Universitaire (CHU) of Orléans for approval prior to implementation. Any changes to the protocol will be reported in the publications resulting from this study.

In the unlikely event of study termination, the reasons for termination will be documented, and all data collected up to that point will be handled in accordance with ethics guidelines and data protection regulations.

## Data Availability

No datasets were generated or analysed during the current study. All relevant data from this study will be made available upon study completion.

## IV. Authors’ contributions

W.K. wrote the study protocol submitted for publication, developed and finalized the questionnaire to be completed by participants, and prepared the participant information sheet. R.S. corrected and supervised the writing of the manuscript. Together with R.S., W.K. presented the study to the Directors General of Services of the three regions and to the ethics committee. W.K., together with R.S. and S.O., presented the study and continues to present it to stakeholders, including educational directors, who will assist in conducting the study. W.K. will be responsible for data collection, data analysis, and reporting.

R.S. was responsible for ethical oversight and logistics coordination necessary to obtain authorization to initiate the study. R.S. introduced W.K. into the “Covidor” network [61]. S.O., R.N, S.M-L, P.B., and O.V. reviewed the protocol and the manuscript. All authors approved the submitted version.

## V. Acknowledgments

The authors would first like to thank the Établissement Public de santé Mentale Georges Daumézon (EPSM du Loiret Georges DAUMEZON) for the financial support of the study, as well as the Centre Hospitalier Universitaire (CHU) of Orléans for granting ethical approval for its conduct. They also thank the Centre de Biophysique Moléculaire (CBM), Centre National de la Recherche Scientifique (CNRS), and in particular the Neurobiology of Receptors & Therapeutic Innovations (NEURRIT) team, for their institutional support.

Special thanks are extended to all early childhood professionals who will participate in this study, as well as to Orléans Métropole, the Communauté de Communes des Terres du Val de Loire (CCTVL), and Fleury-les-Aubrais regions for authorizing for the conduct of the study in their regions and for their support in facilitating its implementation and success.

## VI. Supporting Information

S1 File: Participant information letter. This sheet will explain the objectives and procedure as well as state that our study complies with ethical guidelines, inform participants of their rights, and state that by continuing, they do not object to participating

S2 File: The questionnaire. The self-administered online questionnaire used to collect data.

